# Retrieving Lab Test Related Questions from Social Q&A Sites by Combining Shallow Features and Deep Representations

**DOI:** 10.1101/2020.08.08.20170753

**Authors:** Yu Lu, Xiao Luo, Zhan Zhang, Haoran Ding, Zhe He

**Author notes:** Yu Lu was at Pace University when the paper was submitted and at Florida State University when the final version of this paper was submitted. Corresponding author: Zhe He.

## Abstract

Patients face challenges in accurately interpreting their lab test results. To fulfill their knowledge gap, patients often turn to online resources, such as Community Question-Answering (CQA) sites, to seek meaningful information and support from their peers. Retrieving the most relevant information to patients’ queries is important to help patients understand lab test results. However, few studies investigated the retrieval of lab test-related questions on CQA platforms. To address this research gap, we build and evaluate a system that automatically ranks questions about lab tests based on their similarity to a given question. The system is tested using diabetes-related questions collected from Yahoo! Answers’ health section. Experimental results show that the regression-weighted combination of deep representations and shallow features was most effective in the Yahoo! Answers dataset. The proposed system can be extended to medical question retrieval, where questions contain a variety of lab tests.

## Introduction

With the wide adoption of patient-facing technologies, such as patient portals connected to healthcare providers’ electronic health record (EHR) systems, patients now have easy, timely, and direct access to their clinical data. However, studies have shown that many patients, especially those with lower health literacy, have difficulty understanding the abundant data in the patient portals.^1, 2^ Therefore, in order to make sense of the available data for personalized decision-making, patients often turn to online resources to seek information and get help from someone who has the expertise or similar experience.^3^ Among various online platforms, Community Question-Answering (CQA) sites have been one of the most popular channels due to their interactivity and little restriction, and more importantly, it is a good source for people to contextualize their health concerns and identify information that might be relevant to him/her.^4^ Retrieving the most relevant health information and posts on such platforms is crucial as it fills patients’ knowledge gap on lab test results for subsequent decision making.^5–7^

A few studies have proposed computational methods for retrieving similar questions on CQA sites to support patients’ information-seeking.^8–10^ However, the research on similar question retrieval concerning lab tests is limited, despite those questions constitute a significant portion of medical questions asked online. It is important to improve the question retrieval pertaining to lab test results because most patient portals only provide lab test results with a reference range that is not contextualized in the specific patient’s condition.^11^ For example, the reference range of Thyroid-Stimulating Hormone (TSH) for pregnant women is different from the general public due to its correlation to the risk of miscarriage.^12^ If a patient is seeking information about TSH test for pregnant women, it is critical to retrieve relevant questions asked by others who share similar experiences (e.g., those who were pregnant and received similar test results).

To this end, we build and evaluate a system consisting of multiple deep text representations, extracted lab test information, and engineered features to identify the relevant questions about lab test results on CQA platforms. We experiment with different text representations including bag-of-words (BoW), the deep contextualized word embeddings – ELMo, the sentence-level embedding – Universal Sentence Encoder (USE), and the unsupervised language representation model – BERT. The lab test information includes the types of lab tests and the range of the test results. We also investigate the use of shallow features, including sentence length and type of questions. As prior work found that questions and answers in *Yahoo! Answers* have good coverage of UMLS concepts,^13^ in this study, we extract UMLS concepts from the questions and use them as a shallow feature to construct the vector representation of the questions. The system is evaluated using questions posted on Yahoo! Answers’ diabetes category of the health section between 2009 and 2014. Specifically, we evaluate various combinations of deep representations and shallow features using questions that contain three major lab tests related to diabetes: creatinine, HbA1c, and glucose. In order to investigate whether our system degrades the performance on the questions that are not lab test-related, a group of questions without any lab tests is used to evaluate the system as well.

The contribution of our work is two-fold: First, we develop a system that automatically retrieves similar questions to a given question pertaining to lab tests from a large corpus of a Social QA site. Second, we demonstrate the use of both shallow features and deep representations in retrieving similar questions regarding lab tests. To the best of our knowledge, this is the first study that focuses on retrieving lab test-related questions on CQA sites.

## Methods

Figure 1 depicts the proposed system to retrieve similar questions regarding lab tests from CQA sites. A series of data pre-processing steps, including converting all words into lower cases and removing stop words as well as redundant punctuation, was applied to both query questions and candidate questions in a corpus. Then, vector representations of queries and candidates were constructed using BoW and deep-learning-based approaches, such as Embeddings from Language Models (ELMo), Universal Sentence Encoder (USE), and Deep Bidirectional Transformers for Language Understanding (BERT). BoW approaches were employed on all the words in the queries and candidates, and on the UMLS concepts extracted with MetaMap.^14^ Lab tests information was extracted from the question using the ValX tool.^15^ Some engineered features, such as sentence length and type of questions, were also considered to evaluate the similarity between queries and candidates. The similarity of a given query to each candidate was evaluated based on their vector representations, extracted lab test information, and engineered features. All candidates were ranked based on their similarities to the query. To evaluate system performance, we recruited human annotators to evaluate a set of selected query questions and candidate questions, then compared against the ranking generated by the proposed system. The details of each component are described in the following subsections.

**Figure 1.**
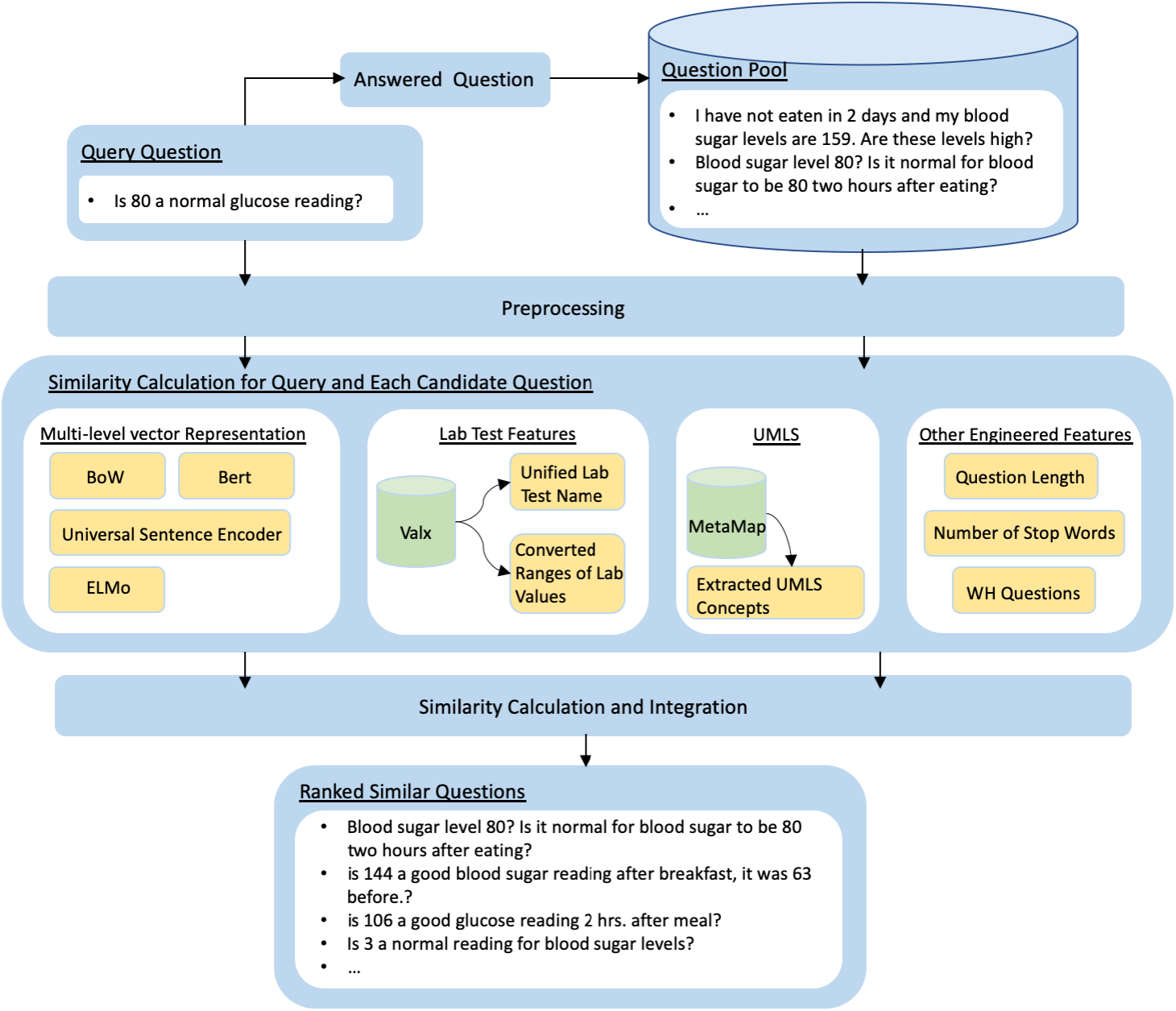
System Overview.

### Dataset and Preprocessing

The dataset we used was collected from the diabetes section of *Yahoo! Answers*, which consists of 58,188 questions posted between 2009 and 2014. We combined the question title and content to preserve the full meaning of questions. Using Valx – a system for extracting numeric lab test comparison statements from the text,^15^ we found that 31,165 questions (53.6%) mention at least one clinical lab results with a value, which shows the importance of building such a system to retrieve medical questions concerning lab tests. We employed Valx to identify and extract lab tests within text content and then convert the extracted lab test values to standardized measurement units. For example, given the text “My glucose is 5.5 mmol/L”, the lab test “glucose” and lab test value “5.5 mmol/L” were extracted. The lab test value was then converted to “100 mg/dl” to make it consistent with other lab results using the unit “mg/dl”.

Analysis of questions’ length distribution shows that most questions have five to twenty words. Hence, in this research, we considered 13,952 questions with five to twenty words. Among these questions, 305 questions contain three diabetes-related lab tests: glucose, HbA1C, and creatinine. To evaluate the robustness of the system, we also randomly selected 2,695 questions without lab tests. In total, 3,000 questions were included for evaluation.

### Traditional and Deep Representations of the Questions

We investigated various representations of the questions, including the BoW/TF-IDF and state-of-the-art deep-learningbased approaches such as ELMo, USE, and BERT. Based on the previous research on biomedical and clinical NLP,^16, 17^ these deep representations can better capture the semantic relationships between the words within the text. The ELMo embedding was trained using the bi-directional Long-Short Term Memory (biLSTM) model, whereas USE and BERT were trained using a transformer architecture. Stemming was performed to the words in the questions when ELMo and USE were used. Since BERT works with tokens, stemming was not applied. Cosine similarity was used to measure the similarity between the query question and candidate questions. When constructing embedding vectors, lab test related words were not and should not be removed.

**Bag of Words (BoW):** The traditional vector space model with TF-IDF^18^ weighting scheme was employed as a basic vector representation. The BoW representation does not consider the semantic relations of the words within the text.

**Embeddings from Language Models (ELMo):** ELMo^19^ derives from the bi-directional language models (biLMs), which take an entire sentence as input. ELMo generates word embeddings by considering the context words surrounding each other. Given a sequence of tokens, a forward language model (LM) computes the probability of the sequence by calculating one token’s probability based on all previous tokens. A backward LM works the same way as the forward LM except it scans through the sequence in reverse order to predict the previous token based on the future context-dependent tokens. The training objective is to maximize the log probability of the forward and backward directions jointly. The output of ELMo is word embeddings, which are the combinations of the intermediate layer representations in the biLM. We used the average of the word embeddings as the sentence embedding. In this research, we used the pre-trained ELMo trained on the 1 Billion Word Benchmark.^20^ The output vector has 3,072 dimensions.

**Universal Sentence Encoder (USE):** Different from ELMo, USE^21^ is a sentence-based embedding model consisting of two components. One generates embedding vectors by adopting the encoding sub-graph of the transformer architecture. The other utilizes the deep-average network, which averages the input embeddings for words and bi-grams and then generates sentence embeddings through passing the average embeddings into a feedforward deep neural network (DNN). In this research, we used the pre-trained USE model. The input to the USE model is a lower-cased PTB tokenized string.^21^ The output is a sentence embedding of a 512-dimensional vector.

**Deep Bidirectional Transformers for Language Understanding (BERT):** The BERT is based on bidirectional self-attention.^22^ Different from other embeddings, such as Word2Vec,^23^ the inputs to the BERT model are not vectors that represent words. Instead, the input includes the tokens, segments, and position embeddings. The token embedding is WordPiece embeddings^24^ that contains 30k tokens. The base BERT model is pre-trained using two unsupervised tasks: (1) Masked Language Model (LM) - a task to predict some random masked tokens in the input. The objective is to train bidirectional encoder. (2) Next Sentence Prediction (NSP) - a task to predict the following sentence of the input sentence. The objective is to understand sentence relationships so that the pre-trained BERT model can be a better fit for other NLP applications, such as Question Answering (QA) and Natural Language Inference (NLI), where sentence relationships are crucial. In this research, we used a pre-trained ClinicalBERT^25^ model trained with clinical notes from MIMIC-III.

### Lab Test Feature Extraction

In this study, we focused on the three lab tests that are most relevant to diabetes diagnosis and management: creatinine, HbA1c, and glucose. Both the type of lab test and the range of the corresponding numeric results were extracted from the questions to measure the similarity between the questions. A numeric expression extraction tool, Valx^15^ was used to extract lab test information from the questions. Valx first extracted numeric values, units (e.g., mmol/l), and comparison operators (e.g., equal to). It then identified lab test variables using hybrid knowledge, including contextual knowledge, domain knowledge, the Unified Medical Language System (UMLS) Metathesaurus.^26^ It also normalized the measurement units, such as ‘mg/dl’, ‘g/l’, ‘mmol/l’. Table 1 presents examples of extracted lab test information using Valx. Each extracted lab has a standardized name, a value, and a measurement unit (highlighted in Table 1).

**Table 1.** Examples of Extracted Lab Tests

| Text | Extracted Lab Tests |
| --- | --- |
| “Fasting blood sugar is 130. am i diabetic?” | Glucose equal to 7.22 mmol/l |
| “is an A1c level higher than 8.0 bad?” | HbA1C greater than 8.0 % |
| “my creatinine equal lower than 0.5. Is it ok?” | creatinine lower than 0.5 mg/dL |

In this study, we first employed Valx to determine whether any of the three lab tests exist in each question. Each of the three lab tests has two features the output of Valx, namely lab type (binary feature) and lab result (categorical feature). If any of these three lab tests are mentioned in a question, the corresponding lab test type feature is 1, otherwise 0. For the value range feature, the extracted lab test results were converted to tertiary ranges. Mayo Clinic websites were used as references for range information of each lab test as shown in Table 2, respectively.^27–29^

**Table 2.** Ranges for The Three Lab Tests

| Lab Test | Range | Indication |
| --- | --- | --- |
| Creatinine | Below 0.84 mg/dL (or below 74.3 mmol/L) | Low |
|  | 0.84 to 1.21 mg/dL (or 74.3 to 107 mmol/L) | Normal |
|  | Above 1.21 mg/dL (or above 107 mmol/L) | High |
| Glucose | Below 100 mg/dL (or below 5.6 mmol/L) | Normal |
|  | 100 to 125 mg/dL (or 5.6 to 6.9 mmol/L) | Pre-diabetic |
|  | Above 126 mg/dL (or above 7 mmol/L) | Diabetic |
| HbA1c | Below 5.7% | Normal |
|  | 5.7% to 6.4% | Pre-diabetic |
|  | 6.5% or above | Diabetic |

We used Equation 1 to measure the similarity between the lab test and test values mentioned in the query and candidate questions, where *y_a_* is the min-max normalized feature value of a query question, and *y_b_* is the min-max normalized feature value of a candidate question.

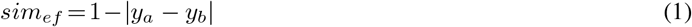

### UMLS Concepts Extraction and Question Representation

We also investigated the effectiveness of integrating biomedical ontologies into our system. Specifically, the UMLS^26^ was used to measure the semantic similarity between questions. The UMLS is a comprehensive thesaurus that consists of over 3 million concepts and over 9 million terms.^26^ Terms with the same meaning are mapped to the same concept. In order to recognize and extract UMLS concepts from question posts, we leveraged MetaMap, a tool for mapping biomedical text to the UMLS metathesaurus.^14^ The MetaMap tool was run on the entire question pool to identify UMLS concepts of each question. Given a text, MetaMap returns the output in a human-readable format, consisting of a score indicating the degree of matching between the phrase and the mapped concept, CUI (Concept Unique Identifier), matched concept, preferred term of the concept, and the semantic type of the concept. The Word Sense Disambiguation (WSD) feature of the MetaMap was used to disambiguate terms. In this research, we included concepts that are assigned a set of diabetes-related semantic types including Physical Object, Substance, Health Care Related Organization, Cell Function, Clinical Attribute, Genetic Function, Organism Attribute, Organ or Tissue Function, Physiologic Function, Biologic Function, Laboratory or Test Result, Diagnostic Procedure, Health Care Activity, Laboratory Procedure, and Therapeutic or Preventive Procedure.

Besides, concepts with a matching score of less than 600 were not considered. The remaining concepts of questions were transformed into vector representations using the TF-IDF weighting scheme. We used cosine similarity to measure the similarity between vectors of the concepts in queries and concepts in candidates.

### Other Engineered Features (EF)

We also investigated other engineered features including the sentence length, the number of stop words (e.g., “the”, “a”, “an”, “in”), and whether it is a WH question (i.e., questions starting with “what”, “how”, “when”, “why”).

### Overall Similarity Calculation and Question Ranking

To calculate the similarity between query and candidate questions, we considered traditional BoW and various deep representations, extracted lab features, a representation based on extracted UMLS concepts, and the additional engineered features through weighted linear combination (Equation 2). The *sim_vf_* corresponds to the similarity measures of shallow and deep representations, *sim_lab___test_* corresponds to the similarity measures based on lab features, *sim_ef_* corresponds to the similarity measures of engineered features, and *sim_umls_* corresponds to the similarity based on the representation using the extracted UMLS features. In this research, the weights were optimized through extensive evaluations. The candidate questions were ranked based on their final values from largest to smallest. The ranked results were then evaluated by human annotators recruited for this research.

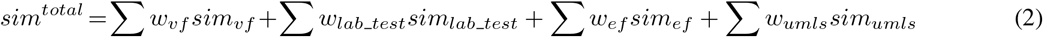

### Linear Combination of Similarity Measures

To optimize the weights for different similarity measure components, we adopted the Linear Regression model from the Weka library^30^ to train a supervised model to learns the weights. For the model, the gold standard generated by the annotators was used for each query-candidate question pair as the dependent variable. The similarity measures are input variables. The regression model learns the weight for each input variable as well as the model deviation. The weighting scheme of the linear regression model for each group of questions is shown in Table 3. The trained model was used to predict the similarity rating of any given query-candidate question pairs.

**Table 3.** Weighting scheme of the linear regression model for each group of questions

| Group | Linear Regression Model |
| --- | --- |
| Creatinine | GS = -2.0886 * ELMo + 2.3099 * USE + 0.9988 * Lab test + 0.8074 * Lab range + 13.7115 * Bert - 11.8422 |
| Glucose | GS = 3.5487 * TF-IDF + 6.3141 * USE + 0.8978 * Lab test - 3.6096 |
| HbA1c | GS = -1.2676 * Sentence length - 0.5579 * WH question type + 1.7152 * Lab test + 12.9366 * Bert - 9.9048 |
| No Lab | GS = 3.3139 * TF-IDF - 1.8626 * ELMo + 1.8375 * USE + 0.6844 * Stopword count + 0.917 |
| Multiple Lab | GS = 3.7002 * TF-IDF - 4.9514 * ELMo + 3.2681 * USE + 1.0927 * Sentence length - 1.3168 * WH question type + 0.5748 * Glucose range + 0.8234 * HbA1c test + 0.8126 * Creatinine test + 2.0664 |

**Table 4.** Pearson correlation results for questions contain lab test

| Method | Creatinine | p Value | Glucose | p Value | HbA1c | p Value | No Lab Tests | p Value | Multi-Lab Tests | p Value |
| --- | --- | --- | --- | --- | --- | --- | --- | --- | --- | --- |
| BoW | -0.010 | 0.925 | 0.300 | 0.002 | -0.107 | 0.290 | **0.489** | **2.47E-07** | 0.592 | 5.84E-06 |
| USE only | 0.170 | 0.092 | **0.608** | **1.93E-11** | 0.314 | 0.001 | **0.379** | **1.02E-04** | 0.530 | 7.64E-05 |
| ELMo only | 0.178 | 0.078 | 0.280 | 0.005 | 0.242 | 0.015 | -0.023 | 0.822 | 0.384 | 6E-03 |
| Bert only | 0.448 | 3.32E-06 | 0.229 | 0.022 | 0.338 | 5.906E-04 | 0.284 | 0.004 | 0.350 | 0.013 |
| BoW+lab | 0.740 | 2.29E-18 | 0.507 | 7.171E-08 | 0.596 | 5.868E-11 | - | - | 0.698 | 1.72E-08 |
| USE+lab | **0.761** | **6.09E-20** | 0.550 | 3.19E-09 | **0.638** | **9.38E-13** | - | - | 0.686 | 3.88E-08 |
| ELMo+lab | 0.726 | 1.76E-17 | 0.488 | 2.62E-07 | 0.612 | 1.28E-11 | - | - | 0.619 | 1.63E-06 |
| Bert+lab | 0.732 | 6.95E-18 | 0.470 | 8.03E-07 | 0.590 | 1.07E-10 | - | - | 0.613 | 2.22E-06 |
| BoW+lab+EF | 0.684 | 5.98E-15 | 0.368 | 1.66E-04 | 0.448 | 2.99E-06 | - | - | 0.400 | 0.004 |
| USE+lab+EF | 0.709 | 2.26E-16 | 0.428 | 8.90E-06 | 0.518 | 3.29E-08 | - | - | 0.395 | 0.005 |
| ELMo+lab+EF | 0.675 | 1.73E-14 | 0.353 | 3.12E-04 | 0.472 | 7.12E-07 | - | - | 0.297 | 0.036 |
| Bert+lab+EF | 0.680 | 9.78E-15 | 0.333 | 7.21E-04 | 0.446 | 3.26E-06 | - | - | 0.270 | 0.058 |
| BoW+lab+UMLS | 0.713 | 1.32E-16 | 0.494 | 1.75E-07 | 0.573 | 4.73E-10 | - | - | 0.694 | 2.32E-08 |
| USE+lab+UMLS | 0.725 | 2.13E-17 | 0.536 | 1.93E-08 | 0.608 | 1.93E-11 | - | - | 0.686 | 3.89E-08 |
| ELMo+lab+UMLS | 0.693 | 1.96E-15 | 0.469 | 8.39E-07 | 0.593 | 7.78E-11 | - | - | 0.636 | 6.92E-07 |
| Bert+lab+UMLS | 0.695 | 1.50E-15 | 0.455 | 1.96E-06 | 0.562 | 1.22E-09 | - | - | 0.637 | 6.56E-07 |
| ISM | 0.207 | 0.038 | **0.624** | **3.92E-12** | 0.303 | 0.002 | 0.340 | 5.53E-04 | 0.670 | 1.04E-07 |
| ISM+EF | 0.244 | 0.015 | 0.264 | 0.008 | 0.131 | 0.194 | 0.299 | 0.003 | 0.084 | 0.564 |
| ISM+lab | **0.752** | **3.21E-19** | 0.596 | 6.00E-11 | **0.648** | **3.27E-13** | - | - | **0.737** | **1.08E-09** |
| ISM+UMLS | 0.210 | 0.037 | 0.515 | 4.15E-08 | 0.310 | 0.002 | 0.343 | 4.863E-04 | 0.634 | 7.70E-07 |
| ISM+lab+EF | 0.674 | 1.99E-14 | 0.481 | 4.18E-07 | 0.515 | 4.23E-08 | - | - | 0.515 | 1.13E-04 |
| ISM+lab+UMLS | 0.733 | 6.90E-18 | 0.575 | 4.10E-10 | 0.637 | 1.08E-12 | - | - | **0.721** | **3.64E-09** |
| LRw | **0.763** | **2.98E-20** | **0.717** | **5.01E-17** | **0.704** | **3.01E-16** | **0.575** | **3.78E-10** | **0.862** | **8.85E-16** |

### Evaluation

Since there is no publicly available dataset for similar lab-test related question retrieval, we created a labelled dataset for this project. The dataset comprises 450 question pairs. This section describes how the 450 labelled dataset was obtained and how the evaluation was designed. There were 45 query questions, including 10 questions for each lab test (glucose, HbA1c, and creatinine), 10 questions that do not contain any lab tests, and 5 questions containing more than one lab test. The reason to include questions without any lab test is to evaluate whether our system can be generalized to general medical question retrieval tasks. For each query question, we used three models, namely ISM (i.e., BoW+ELMo+USE+BERT)+lab+EF, ELMo, and BoW, to obtain different rankings of the candidate questions. As each feature combination retrieved different candidates, we randomly selected the candidates from each quartile of the three rankings to form an annotation set of 1,350 query-candidate pairs. For each pair, the query had 30 candidates in total (ten candidates from each of the three methods), including a small proportion of duplicated candidates.

For the annotation, We recruited three human annotators to rate the relevance of the 1350 query-candidate question pairs. A guideline of relevance review was given to each annotator. Each annotator was asked to give each candidate question a score between 0 to 5, where 5 means “extremely relevant” and 0 means “not relevant at all”. The candidate questions were independently scored by annotators who have general knowledge about diabetes-related lab tests. After the annotation, we selected 10 distinct questions out of the 30 candidates for each query question based on the distribution of the rating to form an evaluation set of 450 query-candidate pairs. We intended to balance different levels of the relevance of the candidate questions to the query questions by including a balanced number of pairs with different scores in the evaluation set.

To assess the degree of agreement of each user’s rating with respect to the other two, we computed Pearson correlation of the scores of each of the three annotators with respect to the average scores of the remaining two raters. The computation was performed on the evaluation set. The correlations between any two raters are 0.640, 0.640, and 0.614, respectively. We found that the relatively low agreement among the three raters was caused by an outlier rating that significantly differs from the other two ratings in some query-candidate pairs. As such, we removed the outlier rating in each query-candidate pair. Then we computed the Pearson correlation between the remaining two ratings, which is 0.953. This correlation is deemed as the upper bound for our system evaluated with this data set.

For each query-candidate pair, the two ratings were averaged to form a gold-standard rating. Then the Pearson correlation coefficient^31^ between the gold-standard rating and the similarity score was computed for evaluation. To assess the strength of the correlation, the guideline proposed by Evans^32^ was used as a reference.

## Results

Table 7 shows the detailed results. Without considering the lab test features, the deep representations – USE, ELMo, and BERT outperformed the baseline BoW across the three groups of questions that contain lab tests. However, BoW still performed better than the deep representation models on questions with no lab tests. We investigated a few questions without lab tests, and found that one possible reason is that TF-IDF weighting scheme provides higher weight to certain words, whereas other deep representations consider the semantic meaning of the whole question. Among all the deep representation models, USE performed consistently better than ELMo across all except for creatinine questions; BERT outperformed ELMo across all except for glucose questions. Such a discrepancy could be explained by the difference in the architecture of the two models: ELMo maintains an embedding size of 3,072 dimensions among all layers, whereas the embedding size of USE is 512 for both the transformer-based encoders and deep averaging network (DAN); the embedding size of BERT is 768 dimensions. In a high dimensional space, the same collection of words (e.g. sugar and glucose) may have a relatively lower cosine similarity as compared to a lower-dimensional space.^33^ Also, if any of the single representations negatively correlated with the gold-standard rating, the correlation for ISM and the gold-standard would be weakened. As a result, ISM, the integration of the deep representation models and the baseline BoW, did not outperform the best single deep representation model in each group except for glucose questions and multiple-test questions. For these two groups, no negative correlations were found for the single representations. Hence, ISM maintained a stronger correlation with the gold standard than the best-performing single representation in these groups.

**Table 5.** Given a creatinine related query question: *What is Serum Chemistry. My Serum Creatinine shows 1.50MG/DL, what that means what precaution should be take?*, the gold-standard ranking and the ranking of each candidate question using three different methods are listed.

| Candidate Question (Annotated score) | Method |  |
| --- | --- | --- |
|  | BERT+Lab | BERT+Lab+EF |
| *my serum creatinine level is 1.42 is it risky? i am alcoholic (3)* | 3 | 8 |
| *My mother aged 45 has only one kidney.creatinine level 4.2,Urea 50,what diet she should take,what medicine? (6)* | 4 | 7 |
| *If creatinine is given as 278 what is the unit it is expressed? My BUN level is 21. (8)* | 8 | 2 |

**Table 6.** Given a glucose related query question: *Is 80 a normal glucose reading?*, the gold-standard ranking and the ranking of each candidate question using three different methods are listed.

| Candidate Question (Annotated score) | Method |  |
| --- | --- | --- |
|  | ISM | ISM+UMLS |
| *is 144 a good blood glucose reading after breakfast, it was 63 before.? (2)* | 4 | 3 |
| *is 3 a normal reading for blood sugar levels? (4)* | 1 | 1 |
| *is a 13.8 reading on my glucose machine bad? as I am normal at 7 (6)* | 2 | 2 |

**Table 7.**
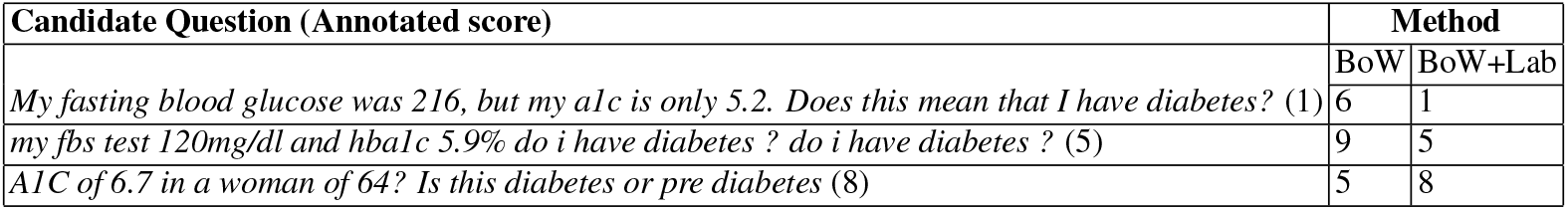
Given a HbA1C related query question: *my a1c is 5.4 do i have pre-diabetes? i am anemic and my a1c is 5.4 do i have pre-diabetes*, the gold-standard ranking and the ranking of each candidate question using three different methods are listed.

The upper-bound correlation was 0.953 according to the correlations between human annotators, which can be deemed as the performance of a typical human. We achieved a high correlation of 0.862 on the multi-lab group with LRw.

## Discussion

In this study, we explored various deep vector representations along with lab test features (the type of lab test and its range), UMLS concepts, and other engineered features, for retrieving similar medical questions regarding lab tests.

In particular, human annotation had a high impact on the evaluation results. Nevertheless, annotation involves uncertainties and challenges. For example, despite a guideline for annotation was introduced to the human annotators, the annotation results may not always align with the rating criteria of each score range in the guideline. Additionally, annotators expressed the feeling that the annotation tends to be subjective even though the guideline was clearly understood. On the other hand, if more detail was provided in the guideline, the annotator concerned that they would have been confused about which score to assign to a specific pair, resulting in ratings that were inconsistent with the criteria provided in the guideline.

### Use of Lab Features

Our results show that including lab features significantly improved the correlation between each method and the goldstandard rating, which aligns with our hypothesis – the integration of lab test related features can effectively enhance the retrieval of similar medical questions that contain clinical lab results. For example, as shown in Table 6, the given query question and the three candidates all made the same type of inquiry and provided HbA1c test results, where both the query question and the first candidate question had HbA1c results in the normal range, while the second and third candidate questions had HbA1c results in the diabetic range. As the simple BoW could not capture the ranges of the test results, its ranking significantly differed from the annotation. By integrating the lab features, BoW+Lab improved the ranking of BoW, and the ranking of the three candidates is consistent with the gold-standard. For questions that contain multiple lab tests, integrating lab-related features also significantly improved the performance over the use of each deep representation method.

### Use of UMLS Features

Although introducing the UMLS features did not enhance each model’s correlation with the gold-standard rating, there is room for future improvement. Due to the limitation of the algorithm, the UMLS mapped some synonymous terms such as *blood sugar*, *blood sugar level*, and *fasting blood glucose* to different concepts. For example, as shown in Table 9, MetaMap identified UMLS concept *normal glucose* of the semantic type *Finding* from the query question. It extracted (1) the concept *Blood Glucose* of the semantic type *Organic Chemical* from the first candidate question, (2) the concept *blood sugar levels* of the semantic type *Laboratory or Test Result* from the second candidate, and (3) the concept *glucose* of the semantic type *Biologically Active Substance; Organic Chemical; Pharmacologic Substance* from the third candidate question. As such, MetaMap extracted four different concepts about glucose. Thus the improvement of the similarity between query and candidate questions is limited. Future work can consider higher level concepts or integration of these concepts through some similarity measurement.

**Table 8.** No Lab: Candidate Questions of Given a query *how can type 1 diabetes be prevented?* that has no lab tests, gold-standard ranking and the ranking of each candidate question using three different methods are listed.

| Candidate Question (Annotated score) | Method |  |
| --- | --- | --- |
|  | ISM | ISM+UMLS |
| *I am really scared i might have type 1 diabetes how can i prevent it????* (1) | 4 | 2 |
| *How can i prevent diabetes?* (2) | 1 | 1 |
| *does type 2 diabetes have a cure?* (6) | 3 | 7 |

**Table 9.**
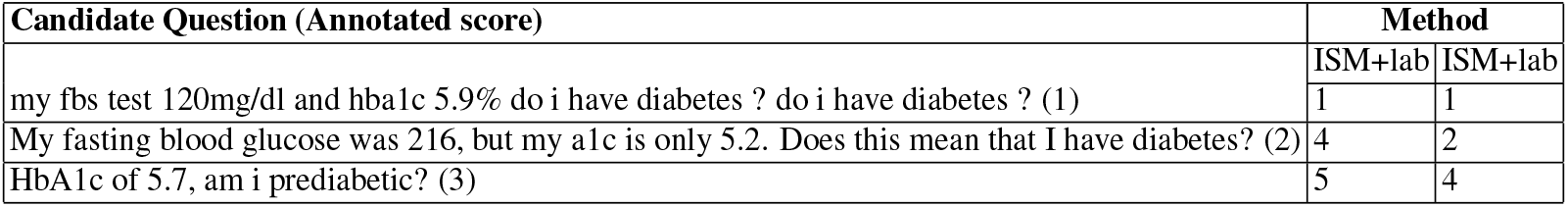
Multiple Lab: Given a query *My fbs is now 5.68mol with hba1c of 5.1 is there a need to take diabetic medicine?* that has no lab tests, the gold-standard ranking and the ranking of each candidate question using three different methods are listed.

Nonetheless, there are cases where UMLS concepts improve the correlation between a method and the rating. For example, as shown in Table 8, MetaMap extracted the key concepts *prevent*, *Type I Diabetes*, *Diabetes*, and *Type II Diabetes* from the questions. Comparing to ISM, ISM+UMLS’s ranking is more consistent with the annotation.

### Use of Engineered Features

Also, it appears that including engineered features also had no apparent positive impact on the methods. This may be attributed to the fact that some of the medical questions may not be WH questions. Also, if two candidate questions asked about different topics while both had a *WH* question or similar sentence length, the engineered feature would in turn impair the ranking. For example, as shown in Table 5, both the second and third candidate questions are *what* questions and have more similar sentence length and the number of stop words to the query question than the first candidate. Even though the three questions all have creatinine test results in the same range, the second and third candidates were identified as less similar to the query question than the first candidate according to the annotation.

### Limitations and Future Work

This study had several limitations: The extraction of glucose results by Valx was not as accurate as creatinine and HbA1c tests. Such errors could impact our results. The adoption of the UMLS features was not effective in optimizing the ranking of candidates, which could be possibly improved by adopting concept similarity measures. As for the human annotation, all the annotators only have basic knowledge of diabetes, which may have introduced disagreements. In future work, we will apply learning algorithms such as polynomial regression to optimize the feature weights. Further analyses and systematic approaches will be applied to evaluate which features contribute more or less to similarity measures. We will also employ multi-layer neural networks for feature learning, and try other data sets.

## Conclusion

This study investigated the combined use of various deep representation models and shallow features in retrieving similar medical questions that contain lab tests. We developed a set of query-candidate question pairs for evaluation. The results show that the regression-weight model outperforms the baseline BoW and the other methods. According to the evaluations by the human annotators, our method was shown to effectively identify the most similar medical questions in *Yahoo! Answers*.

## Data Availability

The data are available upon request.

## Acknowledgements

This study was partially supported by the National Institute on Aging under Award Number R21AG061431; and in part by the National Center for Advancing Translational Sciences under Award Number UL1TR001427. The content is solely the responsibility of the authors and does not necessarily represent the official views of the NIH. We would also like to thank Dr. Sanghee Oh for sharing with us her collected data from Yahoo!Answers.

